# Exploring the relationship between social determinants of health and pediatric injury outcomes at a Northern Tanzania tertiary referral hospital

**DOI:** 10.1101/2025.09.09.25335474

**Authors:** Natalie J Tedford, Modesta Mitao, Timothy Antipas Peter, Linda Minja, Gabriele Nascimento de Oliveira, Raven Mingo, Getrude Nkini, Catherine Staton, Joao R.N. Vissoci, Blandina T. Mmbaga, Elizabeth M. Keating

## Abstract

**Background:** Trauma is a leading cause of death and disability in children globally, with social determinants of health (SDH) affecting health outcomes. Limited data is available on the influence of SDH on outcomes in injured children in low- and middle-income countries (LMICs).

**Objective:** To explore the relationship of SDH factors with morbidity and mortality in pediatric trauma patients at a referral hospital in an LMIC.

**Methods:** This cross-sectional study utilized a prospective pediatric trauma registry at a tertiary hospital in northern Tanzania. It enrolled children under the age of 18 who presented with acute injuries. SDH factors included community type, payment method, insurance status, household composition, daily nutrition, and transfer status. Morbidity was assessed using the Glasgow Outcome Scale Extended Peds (GOS-E Peds). The primary outcomes were mortality and morbidity. Chi-square analysis, logistic regression, and modified Poisson regression modeling were employed to analyze associations between SDH burden and patient-specific outcomes.

**Results:** From November 2020 to January 2024, 877 patients were enrolled, resulting in a mortality rate of 7.0% and 38.8% experiencing poor morbidity outcomes (GOS-E Peds ≥3). Older patients, uninsured patients, those living outside of Moshi Urban, those transferred by ambulance to KCMC, and those sustaining burn injuries had higher odds of mortality in univariable analysis. In adjusted Poisson regression, older patients demonstrated higher odds of morbidity, while those transferred without an ambulance to KCMC showed lower odds of morbidity. Food insecurity emerged as a significant factor influencing survival and poor outcomes, as illustrated in a Sankey diagram (Figure 1) that depicts the pathways to good (GOS-E Peds ≤2) and poor (GOS-E Peds ≥3) outcomes.

**Conclusions:** Several SDH factors, including insurance status and food insecurity, were associated with increased mortality and morbidity. These findings underscore the need for SDH monitoring and targeted interventions to address disparities in pediatric trauma outcomes in LMICs.

## Introduction

Injuries are a leading cause of death in children aged 5-14 years worldwide(1). More than 95% of pediatric injury-related deaths occur in low- and middle-income countries (LMICs)(1), with children in sub-Saharan Africa even more disproportionately affected(2, 3). In LMICs, children are at a higher risk of experiencing injuries due to various influences, including inadequate infrastructure and regulations, lack of specialized medical services, and social factors (i.e., education level, poverty, limited access to health care, inadequate safety measures, etc.). These can significantly affect the incidence and severity of injuries and negatively affect the recovery and health outcomes of affected children(2-5). However, there is a lack of data on the relationship between social determinants of health (SDH) and the health outcomes of injured children in LMICs.

SDH are the circumstances in which people are born, grow, live, work, and age(6). The role of SDH on the health outcomes of children experiencing injuries from trauma in LMICs has been recognized by researchers and policymakers(7-11). There is adequate literature demonstrating that childhood disadvantage is linked to subsequent physical morbidity among adults(7, 12-16). Yet, the literature on SDH and pediatric trauma in LMICs, specifically sub-Saharan African countries, is limited. Furthermore, LMICs often have insufficient integration of health inequality monitoring into their health information systems(17). Understanding the complex interplay between SDH and the health outcomes of injured children can help identify effective strategies to prevent injuries, improve access to care, and enhance the recovery and rehabilitation of affected children.

Given the limited data on pediatric injuries and the impact of SDH on child health and welfare, this exploratory study aimed to identify and investigate the associations of social influencers with the pediatric injury population in Northern Tanzania. To accomplish this, we utilized the SDH domains (economic stability, neighborhood, physical environment, food, community, social context, education, and health care system)(18) to collect various SDH factors for each injured child in our cohort to understand the presence and context of SDH on pediatric injury health outcomes. The findings can inform locally relevant, targeted interventions to address inequalities and subsequently reduce morbidity and mortality from pediatric injuries in this region and other similar settings.

## Materials and Methods

### Study design

This cross-sectional study used data from a prospective pediatric trauma registry at a tertiary referral hospital in Northern Tanzania. Patients were enrolled in the registry upon presentation to the Emergency Department if they were under 18 years old and presented with an acute injury. The trauma registry data collected has been previously described(19). Additional information regarding the trauma registry data collected is included in the Supporting Information (S1 Registry Intake), with SDH factors highlighted. Morbidity was measured using the Glasgow Outcome Scale Extended Peds (GOS-E Peds, Table 1)(20). Outcomes of interest were mortality and morbidity.

**Table 1.**
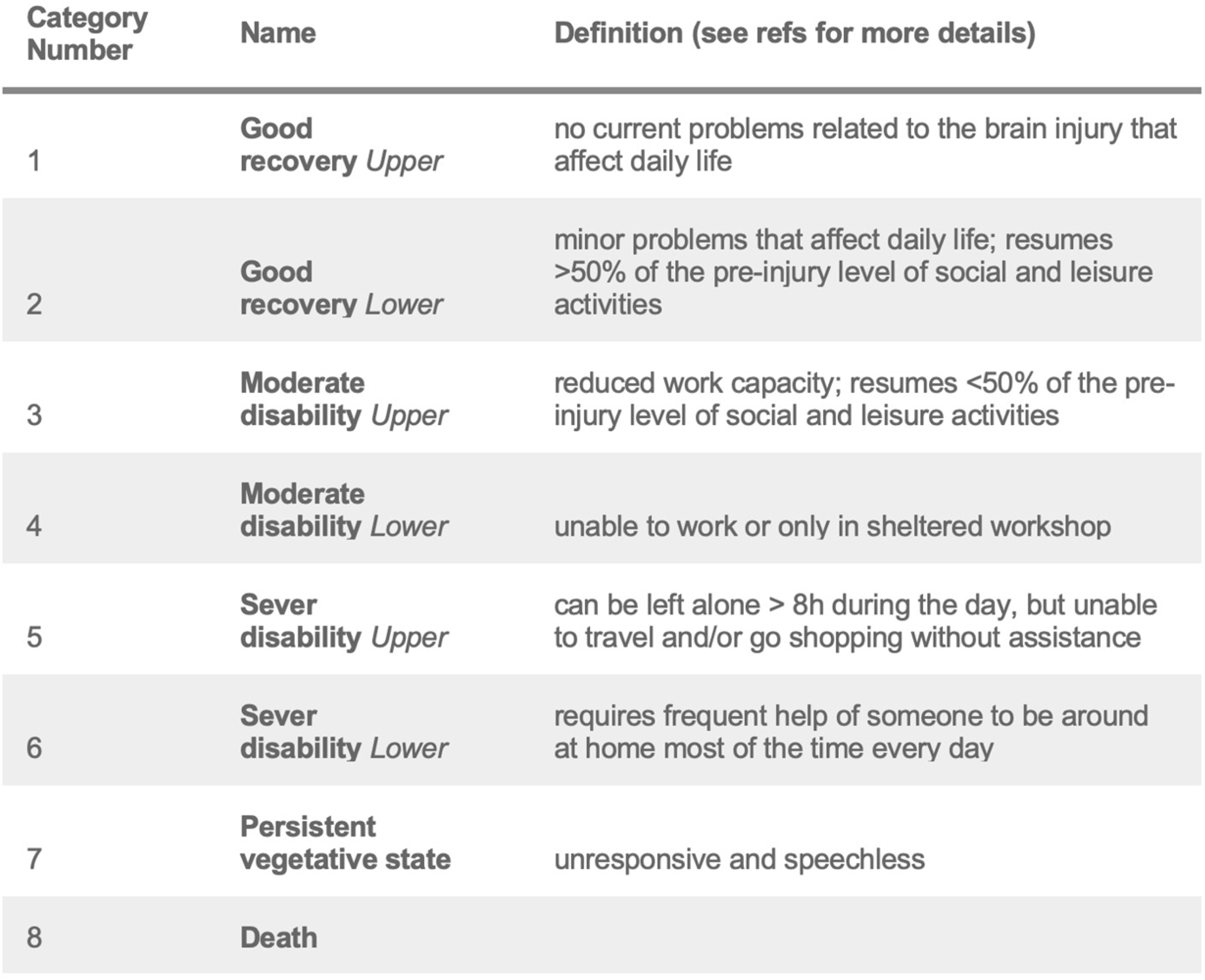
Glasgow Outcome Scale Extended Pediatric (GOS-E Peds) Score Categorizations.

| Category Number | Name | Definition (see refs for more details) |
| --- | --- | --- |
| 1 | **Good recovery** *Upper* | no current problems related to the brain injury that affect daily life |
| 2 | **Good recovery** *Lower* | minor problems that affect daily life; resumes >50% of the pre-injury level of social and leisure activities |
| 3 | **Moderate disability** *Upper* | reduced work capacity; resumes <50% of the pre-injury level of social and leisure activities |
| 4 | **Moderate disability** *Lower* | unable to work or only in sheltered workshop |
| 5 | **Sever disability** *Upper* | can be left alone > 8h during the day, but unable to travel and/or go shopping without assistance |
| 6 | **Sever disability** *Lower* | requires frequent help of someone to be around at home most of the time every day |
| 7 | **Persistent vegetative state** | unresponsive and speechless |
| 8 | **Death** |  |

This study and all procedures received ethical approval from the institutional review boards at the Tanzanian National Institute for Medical Research, Kilimanjaro Christian Medical Centre, and the University of Utah. The ethics committees waived the need for consent.

### Inclusivity in global research

The supporting information (S2 Checklist) includes additional information regarding the ethical, cultural, and scientific considerations specific to inclusivity in global research.

### Study setting

The study occurred at Kilimanjaro Christian Medical Centre (KCMC), a zonal referral hospital for the northern regions of Tanzania (Arusha, Dodoma, Kilimanjaro, Singida, and Tanga). KCMC is one of four zonal referral hospitals in Tanzania and has a catchment population of 12,000,000 persons. KCMC is in the Kilimanjaro region of Northern Tanzania and is a referral site for children with injuries requiring specialty investigations, imaging, and treatment. The Emergency Medical Department (EMD) at KCMC sees approximately 1400–1700 pediatric patients annually. KCMC has general surgeons and orthopedic surgeons who treat trauma patients, including children. In addition, KCMC has pediatricians who assist in caring for pediatric trauma patients. There is a specialized burn center at KCMC. There is no formal emergency medical system in Tanzania.

### Participants

The KCMC pediatric trauma registry was launched in November 2020 to define areas for quality improvement in the care of pediatric trauma patients. Patients were enrolled in the registry on presentation to the KCMC EMD if they were less than 18 years old and presented with an injury. Recruitment started on November 1, 2020, and ended on January 31, 2024. As defined by the World Health Organization, injuries are caused by acute exposure to physical agents such as mechanical energy, heat, electricity, chemicals, and ionizing radiation interacting with the body in amounts that exceed the threshold of human tolerance(21). This registry enrolled patients who were injured with fractures, burns, lacerations, traumatic brain injuries, ingestions/poisonings, animal envenomation, road traffic injuries, falls, drownings, penetrating trauma, non-accidental trauma, and others. Registry inclusion criteria included pediatric patients seeking care for any injury that occurred in the last month who survived an evaluation in the EMD. Patients were excluded from the registry if they presented for injury follow-up care.

### Quantitative procedures

The pediatric trauma registry prospectively enrolled all patients less than 18 years of age presenting to the KCMC EMD for treatment of an injury. All registry data were recorded on tablets in REDCap,(22) and the quality of all entries was reviewed by the Principal Investigator (EMK). For this analysis, variables were extracted from the REDCap® database, including demographics, mid-upper arm circumference (MUAC), the community where the patient lives, caregiver education and employment, the form of payment for healthcare services, food insecurity, mechanism of injury, time of injury, transfer status to KCMC, and patient outcomes including in-hospital mortality and morbidity. Patients with missing outcomes were excluded. Continuous data were represented as means with standard deviation or medians with interquartile ranges (IQR). We used Chi-squared tests to compare survivors and non-survivors across key SDH-related demographics and characteristics. All analyses were performed in R Statistical software (version 2023.12.1+402).

### Statistical analysis

Statistical analysis was performed using R statistical software (version 2023.12.1+402). Categorical variables were presented as absolute numbers (n) and relative (%) frequencies. Continuous variables were expressed as median and IQR (Interquartile range). The Chi-square and Wilcoxon rank sum tests were used to test the association between patients’ sociodemographic characteristics and outcomes.

The Akaike Information Criteria (AIC) was used for model selection. The model with the lowest AIC was selected from an ordinary logistic regression model compared with a modified Poisson model.

Univariable and multivariable logistic regression models were performed to study factors associated with survivor status (Survivor/Non-survivor), and adjustment for potential confounders was performed in the multivariable model. Statistical significance was defined as p<0.05. Assumptions were checked, including multicollinearity, linearity, and influential values. A modified Poisson regression model with standard errors was estimated to study factors associated with morbidity (GOS-E Peds scores ≥3). Assumptions were checked, and appropriate measures were taken to prevent violations. A Sankey diagram was plotted to show the flow of SDH to the patient’s outcome. A complete case analysis was done, and missing data was deemed missing at random.

## Results

From November 2020 to January 2024, 877 patients were enrolled. The majority were males (n=554, 63%), and the median age was 7.0 (IQR 4-12) years (Table 2a). Most patients were not insured (n=694, 79%), resided in Moshi Urban (n=251, 29%), lived with both parents (n=541, 62%), had parents with no formal education (n=392, 45%), arrived by ambulance to KCMC (n=473, 54%), and had adequate nutritional status per median upper arm circumference (MUAC) measurements (18.2 cm, IQR 17.0-21.0) (Table 2). Most families did not require a social worker consultation (n=782, 89%) and reported being food secure (n=537, 61%). The mortality rate was 7.0%, with burn injuries representing the highest burden of injury resulting in mortality (n=25, 41%) (Table 2a). The morbidity with a poor outcome (GOS-E Peds ≥3) was 38.8% (Table 2b).

**Table 2a:** Socio-demographic characteristics of participants stratified by survivor status.

| Characteristic | Overall<br>N = 877<sup>1</sup> | Non-Survivors<br>N = 61<sup>1</sup> | Survivors<br>N = 816<sup>1</sup> | p-value<sup>2</sup> |
| --- | --- | --- | --- | --- |
| **Age (Yrs) – Median (IQR)** | 7.0 (4.0, 12.0) | 7.0 (3.0, 12.0) | 7.0 (4.0, 12.0) | 0.818 |
| Unknown | 34 | 4 | 30 |  |
| **Sex** |  |  |  | 0.470 |
| Male | 554 (63.3%) | 36 (59.0%) | 518 (63.6%) |  |
| Female | 321 (36.7%) | 25 (41.0%) | 296 (36.4%) |  |
| Unknown | 2 | 0 | 2 |  |
| **Insurance** |  |  |  | 0.012* |
| Insured | 183 (20.9%) | 5 (8.2%) | 178 (21.8%) |  |
| Not insured | 694 (79.1%) | 56 (91.8%) | 638 (78.2%) |  |
| **Residence** |  |  |  | 0.015* |
| Moshi Urban | 251 (28.7%) | 8 (13.3%) | 243 (29.8%) |  |
| Moshi Rural | 227 (25.9%) | 22 (36.7%) | 205 (25.1%) |  |
| Other | 398 (45.4%) | 30 (50.0%) | 368 (45.1%) |  |
| Unknown | 1 | 1 | 0 |  |
| **Live with** |  |  |  | 0.525 |
| Both Parents | 541 (61.8%) | 33 (55.0%) | 508 (62.3%) |  |
| Others | 145 (16.6%) | 12 (20.0%) | 133 (16.3%) |  |
| Single Parent | 189 (21.6%) | 15 (25.0%) | 174 (21.3%) |  |
| Unknown | 2 | 1 | 1 |  |
| **Education** |  |  |  | 0.570 |
| None | 392 (45.1%) | 31 (51.7%) | 361 (44.6%) |  |
| Primary education | 376 (43.3%) | 23 (38.3%) | 353 (43.6%) |  |
| Secondary & Above Education | 101 (11.6%) | 6 (10.0%) | 95 (11.7%) |  |
| Unknown | 8 | 1 | 7 |  |
| **Transfer Status** |  |  |  | <0.001* |
| Transfer with ambulance (REF) | 473 (54.0%) | 48 (80.0%) | 425 (52.1%) |  |
| Transfer without ambulance | 241 (27.5%) | 8 (13.3%) | 233 (28.6%) |  |
| Seen first at KCMC | 162 (18.5%) | 4 (6.7%) | 158 (19.4%) |  |
| Unknown | 1 | 0 | 1 |  |
| **MUAC (cm) – Median (IQR)** | 18.2 (17.0, 21.0) | 17.2 (16.4, 20.2) | 18.5 (17.0, 21.0) | 0.247 |
| Unknown | 63 | 29 | 34 |  |
| **Social Worker** |  |  |  | 0.307 |
| No | 782 (89.2%) | 52 (85.2%) | 730 (89.5%) |  |
| Yes | 95 (10.8%) | 9 (14.8%) | 86 (10.5%) |  |
| **Food Insecurity** |  |  |  | 0.860 |
| Yes | 340 (38.8%) | 23 (37.7%) | 317 (38.8%) |  |
| No | 537 (61.2%) | 38 (62.3%) | 499 (61.2%) |  |
| **Tribe of caregiver** |  |  |  | 0.990 |
| Chaga | 349 (39.9%) | 24 (40.7%) | 325 (39.8%) |  |
| Others | 393 (44.9%) | 26 (44.1%) | 367 (45.0%) |  |
| Pare | 133 (15.2%) | 9 (15.3%) | 124 (15.2%) |  |
| Unknown | 2 | 2 | 0 |  |
| **Mechanism of Injury** |  |  |  | <0.001* |
| Burn | 100 (11.4%) | 25 (41.0%) | 75 (9.2%) |  |
| Fall | 298 (34.0%) | 9 (14.8%) | 289 (35.5%) |  |
| Others | 184 (21.0%) | 12 (19.7%) | 172 (21.1%) |  |
| Road Traffic Injury | 294 (33.6%) | 15 (24.6%) | 279 (34.2%) |  |
| Unknown | 1 | 0 | 1 |  |

<sup>1</sup>Median (IQR); n (%)<sup>2</sup>Wilcoxon rank sum test; Pearson's Chi-squared test
\*Indicates statistical significance

**Table 2b:**
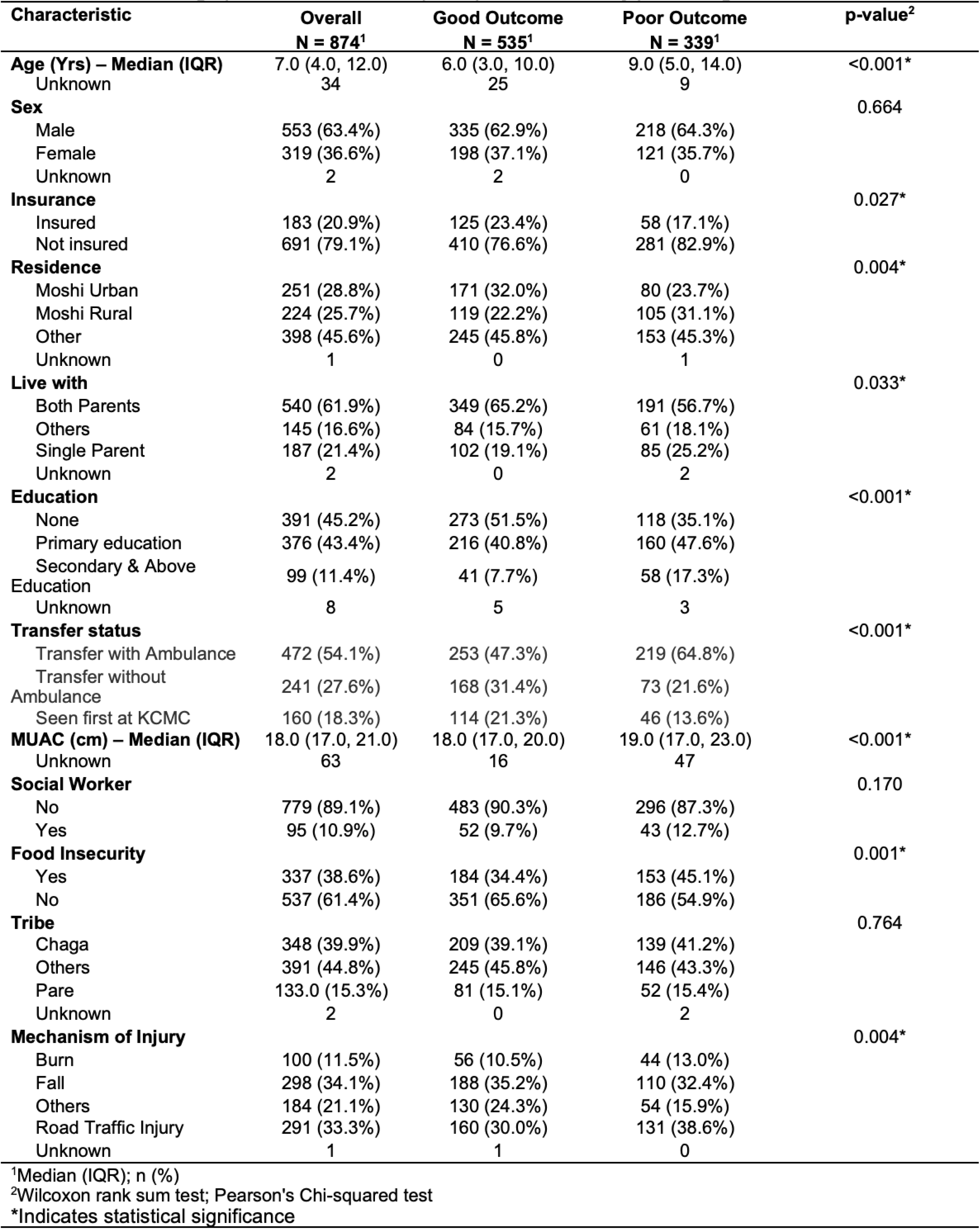
Socio-demographic characteristics of participants stratified by poor and good outcome.

| Characteristic | Overall<br>N = 874<sup>1</sup> | Good Outcome<br>N = 535<sup>1</sup> | Poor Outcome<br>N = 339<sup>1</sup> | p-value<sup>2</sup> |
| --- | --- | --- | --- | --- |
| **Age (Yrs) – Median (IQR)** | 7.0 (4.0, 12.0) | 6.0 (3.0, 10.0) | 9.0 (5.0, 14.0) | <0.001* |
| Unknown | 34 | 25 | 9 |  |
| **Sex** |  |  |  | 0.664 |
| Male | 553 (63.4%) | 335 (62.9%) | 218 (64.3%) |  |
| Female | 319 (36.6%) | 198 (37.1%) | 121 (35.7%) |  |
| Unknown | 2 | 2 | 0 |  |
| **Insurance** |  |  |  | 0.027* |
| Insured | 183 (20.9%) | 125 (23.4%) | 58 (17.1%) |  |
| Not insured | 691 (79.1%) | 410 (76.6%) | 281 (82.9%) |  |
| **Residence** |  |  |  | 0.004* |
| Moshi Urban | 251 (28.8%) | 171 (32.0%) | 80 (23.7%) |  |
| Moshi Rural | 224 (25.7%) | 119 (22.2%) | 105 (31.1%) |  |
| Other | 398 (45.6%) | 245 (45.8%) | 153 (45.3%) |  |
| Unknown | 1 | 0 | 1 |  |
| **Live with** |  |  |  | 0.033* |
| Both Parents | 540 (61.9%) | 349 (65.2%) | 191 (56.7%) |  |
| Others | 145 (16.6%) | 84 (15.7%) | 61 (18.1%) |  |
| Single Parent | 187 (21.4%) | 102 (19.1%) | 85 (25.2%) |  |
| Unknown | 2 | 0 | 2 |  |
| **Education** |  |  |  | <0.001* |
| None | 391 (45.2%) | 273 (51.5%) | 118 (35.1%) |  |
| Primary education | 376 (43.4%) | 216 (40.8%) | 160 (47.6%) |  |
| Secondary & Above | 99 (11.4%) | 41 (7.7%) | 58 (17.3%) |  |
| Unknown | 8 | 5 | 3 |  |
| **Transfer status** |  |  |  | <0.001* |
| Transfer with Ambulance | 472 (54.1%) | 253 (47.3%) | 219 (64.8%) |  |
| Transfer without Ambulance | 241 (27.6%) | 168 (31.4%) | 73 (21.6%) |  |
| Seen first at KCMC | 160 (18.3%) | 114 (21.3%) | 46 (13.6%) |  |
| **MUAC (cm) – Median (IQR)** | 18.0 (17.0, 21.0) | 18.0 (17.0, 20.0) | 19.0 (17.0, 23.0) | <0.001* |
| Unknown | 63 | 16 | 47 |  |
| **Social Worker** |  |  |  | 0.170 |
| No | 779 (89.1%) | 483 (90.3%) | 296 (87.3%) |  |
| Yes | 95 (10.9%) | 52 (9.7%) | 43 (12.7%) |  |
| **Food Insecurity** |  |  |  | 0.001* |
| Yes | 337 (38.6%) | 184 (34.4%) | 153 (45.1%) |  |
| No | 537 (61.4%) | 351 (65.6%) | 186 (54.9%) |  |
| **Tribe** |  |  |  | 0.764 |
| Chaga | 348 (39.9%) | 209 (39.1%) | 139 (41.2%) |  |
| Others | 391 (44.8%) | 245 (45.8%) | 146 (43.3%) |  |
| Pare | 133.0 (15.3%) | 81 (15.1%) | 52 (15.4%) |  |
| Unknown | 2 | 0 | 2 |  |
| **Mechanism of Injury** |  |  |  | 0.004* |
| Burn | 100 (11.5%) | 56 (10.5%) | 44 (13.0%) |  |
| Fall | 298 (34.1%) | 188 (35.2%) | 110 (32.4%) |  |
| Others | 184 (21.1%) | 130 (24.3%) | 54 (15.9%) |  |
| Road Traffic Injury | 291 (33.3%) | 160 (30.0%) | 131 (38.6%) |  |
| Unknown | 1 | 1 | 0 |  |

<sup>1</sup>Median (IQR); n (%)<sup>2</sup>Wilcoxon rank sum test; Pearson's Chi-squared test
\*Indicates statistical significance

Findings from the chi-square test suggest that the patient death was statistically significantly associated with health insurance (p=0.027), resident (p=0.015), transfer status (p<0.001), and mechanism of injury (p<0.001). Wilcoxon rank sum test shows a statistically significant association between age and death (p<0.001) (Table 2a).

Chi-square test findings show a statistically significant association between morbidity and age (p<0.001), health insurance (p=0.027), residence (p=0.004), person who patient lives with (p=0.033), parental education (p<0.001), transfer status (p<0.001), food insecurity (p=0.001), and mechanism of injury (p=0.04). Wilcoxon rank sum test suggests that MUAC measurements (cm) were statistically significantly associated with morbidity (p<0.001) (Table 2b).

Table 3 displays the association between social demographic characteristics and odds of mortality for pediatric patients in a crude and adjusted analysis with potential confounders as listed in Table 2a and Table 2b. In a univariable logistic regression (LR) model, the odds of mortality were higher in uninsured patients and patients residing anywhere except Moshi Urban (OR=2.48 95% CI [1.17-5.88], p=0.026); however, in the adjusted model, residence was not statistically significantly associated with mortality. The odds of mortality were lower in patients transferred to KCMC without an ambulance (OR=0.18 95% CI [0.04-0.59], p=0.011) in the adjusted analysis compared to those transferred to KCMC by ambulance. Patients seen first at KCMC were statistically associated with lower odds of mortality (OR=0.22 95% CI [0.07-0.56], p=0.05) in univariable LR, but was not significant in adjusted analysis. Having a fall as a mechanism of injury was statistically significantly associated with lower odds of mortality by 91% compared to having a burn as a mechanism of injury (OR=0.09, 95% CI [0.04-0.20], p<0.001). After adjusting for potential confounders, the mechanism of injury remained statistically significantly linked to mortality. In the adjusted analysis, age was statistically significantly associated with mortality, where an increase of one year in age was associated with a 27%; however, this association was not statistically significant in the crude analysis. An increase of 1 cm in MUAC measurement was statistically associated with a 22% decrease in the odds of mortality in the adjusted analysis (OR=0.78, 95% CI [0.65-0.93], p=0.008). In the crude analysis, MUAC was not linked to mortality. Food insecurity was not statistically significant in the crude analysis but became significant in the adjusted analysis, where children without food insecurity had doubled the odds of mortality compared to those with food insecurity (OR=2.62, 95% CI [1.07-7.42], P=0.048).

**Table 3:**
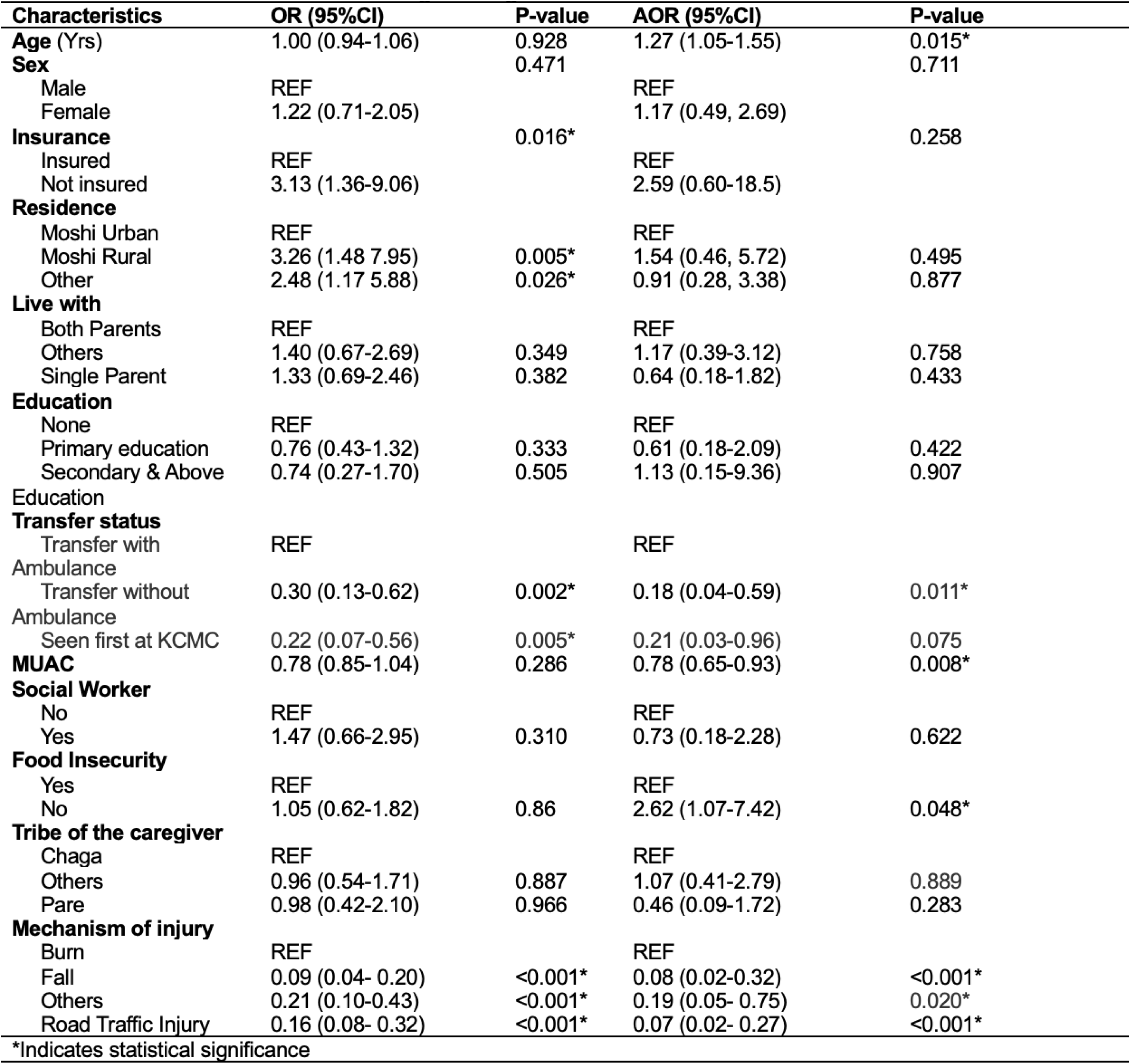
Univariate and multivariate logistic regression models for non-survivors.

In a univariable modified Poisson regression model, the prevalence of morbidity was higher in older patients, those who lived in Moshi Rural areas, those who lived with single parents, those who had a parent with some form of formal education, those who had larger MUAC measurements, and those sustaining injuries other than burns, falls, or RTIs. The prevalence of morbidity was lower in patients who were transferred to KCMC without ambulance transport, seen at KCMC first for presentation, and were food secure at home (Table 4). In the multivariable-adjusted Poisson regression model, the morbidity prevalence was higher in older patients (Table 4). When controlling for the potential confounder of food security, these multivariable-adjusted Poisson regression model results were consistent.

**Table 4:**
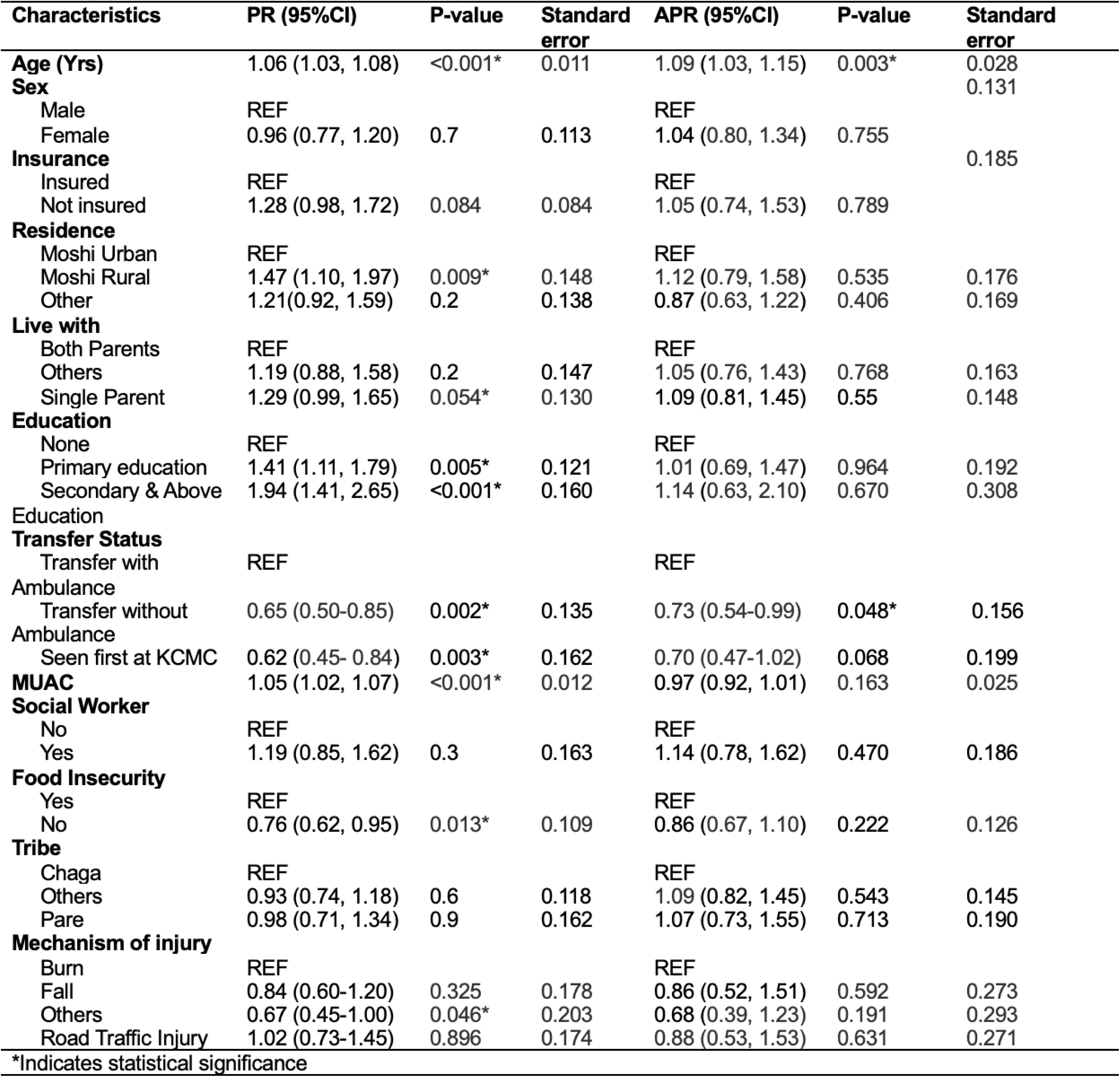
Univariate and multivariate modified Poisson regression models for the prevalence ratio of morbidity/poor outcome (GOS-E Peds ≥3)

Our analysis showed an underlying social vulnerability concept that defined the association with “surviving” and “poor outcomes,” represented by food insecurity. Given the inconsistent findings in our LR modeling and the suspected confounding effect of reported food security, we visualized our data using a Sankey diagram (Figure 1) to better understand the trajectories of good (GOS-E Peds ≤2) and poor (GOS-E Peds ≥3) outcomes for patients to display the social vulnerability associations of food security and insurance status. Figure 1 illustrates the flow of patients through different categories of food security and insurance status, leading to morbidity outcomes. A significant proportion of food-secure patients were uninsured, and most of them had good outcomes (46.9%). Those who experienced poor outcomes among the insured were relatively low (6.6%). Patients with food insecurity had an unbalanced distribution between insured and uninsured status, with the majority (75%) being uninsured. Insured patients with food insecurity appeared to have good outcomes (14.3%), while uninsured patients without food insecurity had higher frequencies of poor outcomes (32.2%), indicating a disparity.

**Figure 1.**
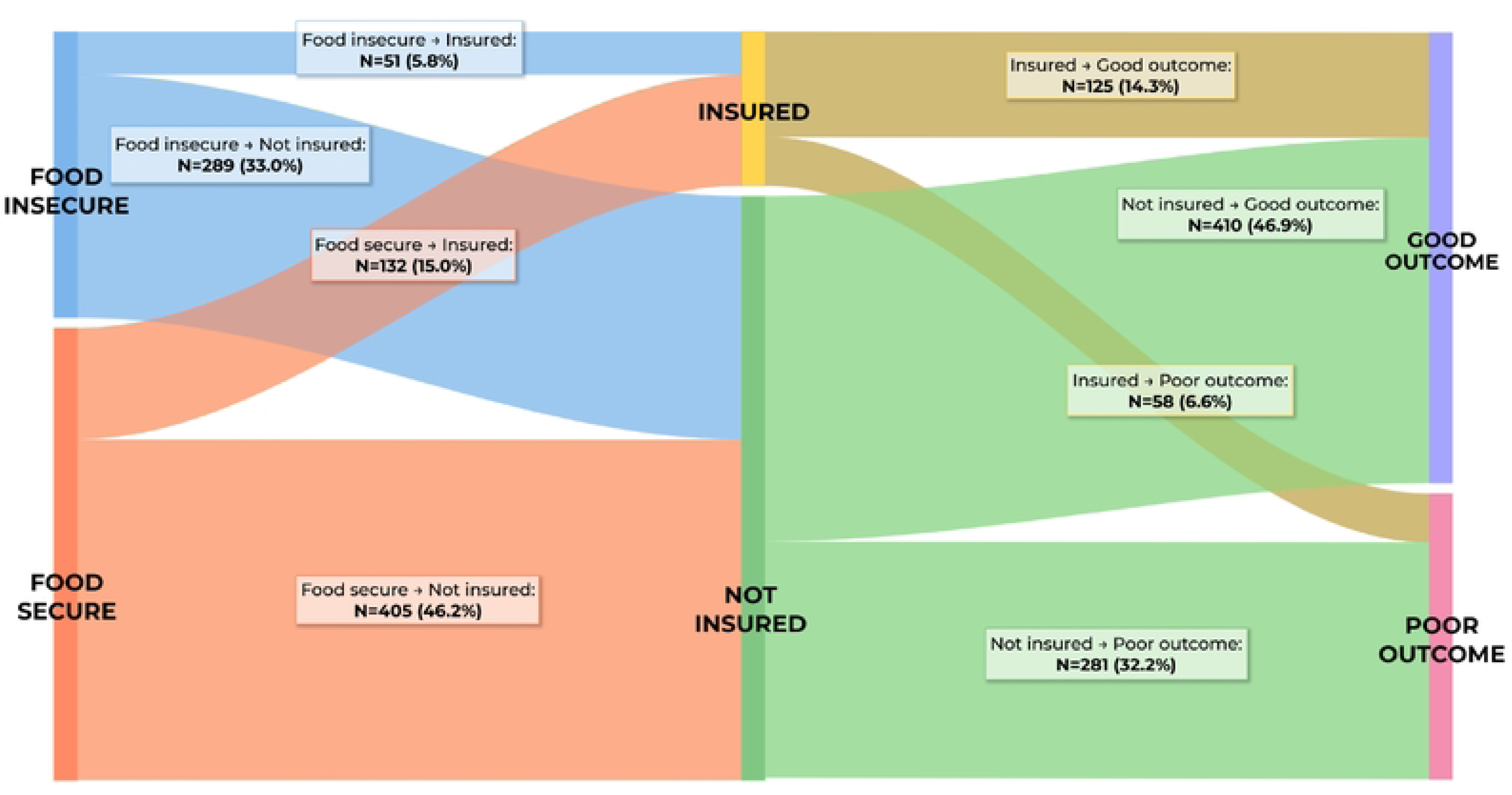
Interconnectedness of Food Insecurity and Insurance Status on Morbidity Description: Sankey Diagram showing the distribution of individuals based on dietary habits (Food secure vs. Food insecure), insurance status (Insured vs. Not Insured), and their corresponding health outcomes (Good vs. Poor). Flow thickness represents the proportion of individuals in each category, highlighting differences in outcome distributions across groups.

## Discussion

In this single-center, cross-sectional analysis of SDH factors in a prospective pediatric trauma registry at a tertiary hospital in Northern Tanzania, we demonstrate SDH influences on morbidity and mortality outcomes of injured children. Our study findings identified associations between the mortality and morbidity of injured children and SDH when related to the patient’s community setting, socioeconomic status, and transportation. Our findings are consistent with reports on the burden of SDH factors associated with an increased risk of injury in LMICs(1, 23-25). This analysis provides insight into factors that could be addressed through in-hospital and community-level interventions.

The relationship between social determinants of health and the health outcomes of children experiencing injuries is influenced by factors such as sex and ethnicity. Sex is an important determinant of injury, with males at a higher risk of injury than females, as our findings demonstrate(26). This may be due to differences in behavior, access to resources, and societal expectations. Ethnicity can also impact the likelihood of injury, as certain ethnic groups may experience higher levels of social exclusion and lack access to resources and services. For example, rural and indigenous populations in many LMICs experience higher rates of injury compared to non-indigenous populations(1).

Our study showed that pediatric injury patients who lacked health insurance had increased mortality and morbidity. This is supported in the literature by other studies in LMICs, which show that insured children have better outcomes(27). In Northern Tanzania, health insurance is for purchase only; thus, only patients who can afford to pay can purchase it. Therefore, it’s possible that health insurance status acts as a proxy for poverty and shows that children with lower socioeconomic status had worse outcomes. This is supported by a study that showed that children from poor households have a higher risk of mortality compared to children from wealthy families(24). Poverty can also influence the quality of healthcare services that children receive following an injury. A study conducted in Pakistan found that children from low-income families were less likely to receive appropriate treatment for injuries compared to children from higher-income families(28). Several studies have highlighted the significant role SDH has on the health outcomes of children experiencing injuries in LMICs, noting that poverty was associated with sustaining more severe injuries, having more extended hospital stays, and experiencing poorer outcomes(4, 13).

Our findings also demonstrate that children transferred to KCMC via ambulance had increased mortality and morbidity. In Northern Tanzania, ambulances do not perform scene transport and are thus mainly for inter-facility transfers. This is a common phenomenon in Sub-Saharan Africa(29). Ambulances are usually prioritized to transport sicker patients, whereas less severe injuries might be transported via public transportation or private cars. Thus, our findings of increased morbidity and mortality in children transported by ambulance are likely since ambulances are transporting children with more severe injuries. A trauma registry study in Malawi supported these findings and found that transport via ambulance was associated with an increased likelihood of admission and more severe injuries(30). Another possible contributor to poorer outcomes in patients transported via ambulance could be a lack of formalized transport care in ambulances. This could also contribute to sicker patients being sicker upon arrival.

In our study, children who lived in Moshi’s urban settings had lower morbidity and mortality than those in rural settings outside of Moshi. This is possibly due to children in these rural settings being at a higher risk of injury due to a lack of access to healthcare and rapid emergency services(31). Access to healthcare is another critical SDH that impacts the health outcomes of children with injuries, and children living in remote and disadvantaged communities have been shown to have limited access to appropriate medical care for injuries, further contributing to poor outcomes(2, 13, 23-25, 31). Additionally, a lack of specialized trauma care centers in LMICs exacerbates the influences of SDH on the health outcomes of children with trauma-related injuries.

In this analysis, food insecurity was associated with increased mortality but not morbidity. The Sankey diagram highlights the critical role of food insecurity and insurance status in determining health outcomes. Patients with food insecurity and lack of insurance were disproportionately affected by poor outcomes. Conversely, insurance coverage appears to mitigate the negative association of food insecurity on health, although not completely. These findings suggest that addressing food insecurity and improving insurance coverage could reduce health disparities and improve morbidity outcomes in vulnerable populations.

The influence of SDH on the health outcomes of children experiencing injuries in LMICs cannot be ignored. Addressing these determinants is crucial to reducing health disparities and improving children’s health outcomes in LMICs. Policies aimed at increasing national health insurance coverage, reducing poverty, improving access to healthcare, and reducing food insecurity can help mitigate the effects of SDH on the health outcomes of children with trauma-related injuries. Such policies should be prioritized in LMICs to ensure that all children have an equal opportunity to access appropriate care and achieve good health outcomes following injuries.

## Limitations

Since this study was based at KCMC and enrolled children who presented for care at KCMC, there is selection and survival bias. We are likely missing some children who presented to outside facilities for care and never made it to KCMC due to the long pre-KCMC timelines that function to pre-select surviving patients. This has the potential to skew the mortality data lower if these children died before arriving at KCMC. In addition, we did not collect quantitative data at the outside health facility level, so we are missing both outcomes and SDH data of patients who were not transferred. Further research is needed at primary levels of healthcare in the system to more accurately understand the drivers or factors influencing the decision to transfer to KCMC and better understand outcomes and SDH barriers. In addition, the lack of a validated food insecurity screening tool created suspected measurement errors since food insecurity was measured with caregiver reports of meals provided. There is potential for social desirability bias influencing this variable as caregivers could have been less comfortable in reporting food insecurity, which could be contributing to our failure to identify a relationship between outcomes and food insecurity.

Further work can be done to develop a culturally sensitive and appropriate validated tool for food insecurity in this population. The SDH factors of our analysis were not the primary focus of the registry’s development, and the tool may not fully capture SDH domains and data. Further, we did not include a variable on injury severity from the registry, which could help benchmark care quality performance with other settings.

## Conclusions

Our study found several SDH factors associated with higher mortality and morbidity in pediatric trauma outcomes. Our findings highlight the importance of understanding SDH-related influences on patient outcomes and integrating inequality monitoring into the health information system in LMICs. Next steps would be interventions that help address and minimize the SDH burden for patients with injuries during hospitalization.

## Data Availability

Yes - all data are fully available without restriction

## Supporting information

**S1 Registry Intake.** Additional information regarding the trauma registry data collected with SDH factors highlighted. (PDF)

**S2 Checklist. Inclusivity in global research.** Additional information regarding the ethical, cultural, and scientific considerations specific to inclusivity in global research.

(DOCX)

## Acknowledgments

The authors would like to acknowledge the pediatric injury patients and their caregivers who participated in our study.

## Author Contributions

**Conceptualization:** Natalie J. Tedford, Modesta Mitao, Timothy Antipas Peter, Linda Minja, Elizabeth M. Keating, Blandina T. Mmbaga.

**Data curation:** Elizabeth M Keating, Gertrude Nkini

**Formal analysis:** Modesta Mitao, Timothy Peter Antipas, Linda Minja, Gabriele Nascimento de Oliveira, Joao RN Vissoci

**Funding acquisition:** Elizabeth M. Keating, Catherine A. Staton, Blandina T. Mmbaga.

**Investigation:** Natalie J. Tedford, Elizabeth M. Keating.

**Methodology:** Natalie J Tedford, Modesta Mitao, Timothy Peter Antipas, Elizabeth M. Keating, Joao RN Vissoci.

**Project administration:** Elizabeth M. Keating, Gertrude Nkini.

**Resources:** Elizabeth M. Keating, Blandina T Mmbaga, Catherine A Staton.

**Software:** Elizabeth M. Keating, Modesta Mitao, Timothy Peter Antipas, Linda Minja, Gabriele Nascimento de Oliveira.

**Supervision:** Elizabeth M. Keating, Blandina T. Mmbaga, Catherine A. Staton, Joao RN Vissoci.

**Visualization:** Gabriele Nascimento de Oliveira, Timothy Peter Antipas

**Writing – original draft:** Natalie J Tedford, Elizabeth M. Keating.

**Writing – review & editing:** Natalie J Tedford, Modesta Mitao, Timothy Peter Antipas, Elizabeth M. Keating, Catherine A. Staton, Linda Minja, Gabriele Nascimento de Oliveira, Raven Mingo, Gertrude Nkini, Joao RN Vissoci, Blandina T. Mmbaga

## References

1. Branche C, Ozanne-Smith J, Oyebite K, Hyder AA. World report on child injury prevention. 2008.

2. Kiragu AW, Dunlop SJ, Mwarumba N, Gidado S, Adesina A, Mwachiro M, et al. Pediatric Trauma Care in Low Resource Settings: Challenges, Opportunities, and Solutions. Frontiers in pediatrics. 2018;6:155-.

3. Abdur-Rahman LO, van As AB, Rode H. Pediatric trauma care in Africa: the evolution and challenges. Semin Pediatr Surg. 2012;21(2):111–5.

4. Saeednejad M, Sadeghian F, Fayaz M, Rafael D, Atlasi R, Kazemzadeh Houjaghan A, et al. Association of Social Determinants of Health and Road Traffic Deaths: A Systematic Review. Bull Emerg Trauma. 2020;8(4):211–7.

5. Ruiz-Casares M. Unintentional childhood injuries in sub-Saharan Africa: an overview of risk and protective factors. J Health Care Poor Underserved. 2009;20(4 Suppl):51-67.

6. WHO Commission on Social Determinants of Health., World Health Organization. Closing the gap in a generation : health equity through action on the social determinants of health : Commission on Social Determinants of Health final report. Geneva, Switzerland: World Health Organization, Commission on Social Determinants of Health; 2008. 246 p. p.

7. Spencer N. Social, economic, and political determinants of child health. Pediatrics. 2003;112(3 Part 2):704-6.

8. Dingake OBK. The Rule of Law as a Social Determinant of Health. Health Hum Rights. 2017;19(2):295–8.

9. Edwards AE, Collins CB. Exploring the influence of social determinants on HIV risk behaviors and the potential application of structural interventions to prevent HIV in women. J Health Dispar Res Pract. 2014;7(SI2):141–55.

10. Lucyk K, McLaren L. Taking stock of the social determinants of health: A scoping review. PLoS One. 2017;12(5):e0177306.

11. Smith J, Griffiths K, Judd J, Crawford G, D’Antoine H, Fisher M, et al. Ten years on from the World Health Organization Commission of Social Determinants of Health: Progress or procrastination? Health Promot J Austr. 2018;29(1):3–7.

12. Goldhagen JL, Shenoda S, Oberg C, Mercer R, Kadir A, Raman S, et al. Rights, justice, and equity: a global agenda for child health and wellbeing. Lancet Child Adolesc Health. 2020;4(1):80–90.

13. Reiner RC, Olsen HE, Ikeda CT, Echko MM, Ballestreros KE, Manguerra H, et al. Diseases, Injuries, and Risk Factors in Child and Adolescent Health, 1990 to 2017: Findings From the Global Burden of Diseases, Injuries, and Risk Factors 2017 Study. JAMA Pediatr. 2019;173(6):e190337.

14. Marmot M, Health CoSDo. Achieving health equity: from root causes to fair outcomes. Lancet. 2007;370(9593):1153-63.

15. Evans GW. Childhood poverty and adult psychological well-being. Proc Natl Acad Sci U S A. 2016;113(52):14949–52.

16. Simkiss DE, Blackburn CM, Mukoro FO, Read JM, Spencer NJ. Childhood disability and socio-economic circumstances in low and middle income countries: systematic review. BMC Pediatr. 2011;11:119.

17. Guerra G, Borde E, Salgado de Snyder VN. Measuring health inequities in low and middle income countries for the development of observatories on inequities and social determinants of health. Int J Equity Health. 2016;15:9.

18. @HIMSS. Social Determinants of Health: @HIMSS; 2021 [updated 2021-02-11. Available from: https://www.himss.org/resources/social-determinants-health.

19. Keating EM, Sakita F, Mmbaga BT, Amiri I, Nkini G, Rent S, et al. Three delays model applied to pediatric injury care seeking in Northern Tanzania: A mixed methods study. PLOS Glob Public Health. 2022;2(8):e0000657.

20. Beers SR, Wisniewski SR, Garcia-Filion P, Tian Y, Hahner T, Berger RP, et al. Validity of a pediatric version of the Glasgow Outcome Scale-Extended. J Neurotrauma. 2012;29(6):1126–39.

21. Holder Y. Injury surveillance guidelines. 2001.

22. Harris PA, Taylor R, Thielke R, Payne J, Gonzalez N, Conde JG. Research electronic data capture (REDCap)—A metadata-driven methodology and workflow process for providing translational research informatics support. Journal of Biomedical Informatics. 2009;42(2):377–81.

23. Haagsma JA, Graetz N, Bolliger I, Naghavi M, Higashi H, Mullany EC, et al. The global burden of injury: incidence, mortality, disability-adjusted life years and time trends from the Global Burden of Disease study 2013. Inj Prev. 2016;22(1):3–18.

24. Birken CS, Macarthur C. Socioeconomic status and injury risk in children. Paediatr Child Health. 2004;9(5):323–5.

25. Bradshaw CJ, Bandi AS, Muktar Z, Hasan MA, Chowdhury TK, Banu T, et al. International Study of the Epidemiology of Paediatric Trauma: PAPSA Research Study. World journal of surgery. 2018;42(6):1885–94.

26. Mock C, Quansah R, Krishnan R, Arreola-Risa C, Rivara F. Strengthening the prevention and care of injuries worldwide. Lancet. 2004;363(9427):2172-9.

27. Mostert S, Njuguna F, van de Ven PM, Olbara G, Kemps LJ, Musimbi J, et al. Influence of health-insurance access and hospital retention policies on childhood cancer treatment in Kenya. Pediatr Blood Cancer. 2014;61(5):913–8.

28. Hyder AA, Razzak JA. The challenges of injuries and trauma in Pakistan: an opportunity for concerted action. Public Health. 2013;127(8):699–703.

29. Mengesha MG, Vella C, Adem EG, Bussa S, Mebrahtu L, Tigneh AY, et al. Use of a trauma registry to drive improvement in the regional trauma network systems in Hawassa, Ethiopia. Eur J Orthop Surg Traumatol. 2023;33(3):541–6.

30. Chokotho LC, Mulwafu W, Nyirenda M, Mbomuwa FJ, Pandit HG, Le G, et al. Establishment of trauma registry at Queen Elizabeth Central Hospital (QECH), Blantyre, Malawi and mapping of high risk geographic areas for trauma. World J Emerg Med. 2019;10(1):33–41.

31. Moshiro R, Furia FF, Massawe A, Mmbaga EJ. Pattern and risk factors for childhood injuries in Dar es Salaam, Tanzania. Afr Health Sci. 2021;21(2):817–25.

